## Supplementary material for "Music and Dance in respiratory disease management in Uganda: A qualitative study of patient and healthcare professional perspectives": Observation sheet

Kupumua Structured Observation Sheet 1

Trial session:

Date:

Observer:

Location:

People present:

Observation of an active session (could be singing, dancing, PR or PR plus music/dance)

| Observation | Session type: |
| --- | --- |
| 1. Body language |  |
| 2. Facial expressions |  |
| 3. Speech/expression |  |
| 4. Interactions between peers |  |
| 5. Interactions with staff |  |
| 6. Physical involvement with music, singing, dancing |  |
| 7. Disease related behaviour (short of breath, coughing, fatigue, resting periods, |  |
| 8. Role within the group. Passive/active. Lead/follow. |  |
| 9. Reflexive researcher responses |  |
