## Supplementary material for "Music and Dance in respiratory disease management in Uganda: A qualitative study of patient and healthcare professional perspectives": Interview guide

### Topic guide interviews –family members

1. How much do you listen to music at home/work?
   a. What type of music?
   b. Who do you listen with?
   c. How do you listen to music (probe for devices/access)
   d. How often?
   e. Do you sing along?
   f. How does music make you feel?
2. Tell us about dancing:
   a. How much do you dance/(other term?) – how often?
   b. What type of dancing?
   c. Who with?
   d. How does it feel to dance?
   e. Does your condition affect your dancing in any way?
   f. What do your friends/family think about that?
3. How is music and dance regarded in your family?
   a. And in your community?
   b. Do people think it is important? Why?
4. What do you think about singing and dancing as a way to improve health for people like your family member?
5. What do you think other people in your community/family would think about that?
6. Would their opinion make any difference to whether your family member might take part?

IF RESPONSES ARE POSTIIVE TO QS 4-6 THEN ASK

7. What do you think could be any difficulties for your family member in doing singing or dance to improve health?
8. What would be the things that might help them?
